# Prenatal Pesticide Exposure and Early Alzheimer Disease-Related Biomarker and Cognitive Changes in Midlife

**DOI:** 10.64898/2026.05.13.26352896

**Authors:** Isha Mhatre-Winters, Piera M. Cirillo, Pam Factor-Litvak, Yoonhee Han, Nickilou Y. Krigbaum, Lauren M. Zimmermann, Bruce G. Link, Young-Mi Go, Dean P. Jones, Barbara A. Cohn, Jason R. Richardson

**Author notes:** **Corresponding Author:** Jason R. Richardson, PhD, Isakson Center for Neurological Disease Research, Department of Physiology and Pharmacology, College of Veterinary Medicine, University of Georgia, Athens, GA 30602, USA.

## Abstract

**Importance:** Alzheimer disease (AD) pathogenesis begins decades before clinical symptoms, yet environmental determinants of early disease risk, particularly during fetal development, remain largely uncharacterized. Prenatal exposure to dichlorodiphenyldichloroethylene (DDE), the primary persistent metabolite of DDT, is a biologically plausible early contributor to AD risk given its long half-life in human tissue and higher levels observed in AD patients. However, prospective human evidence linking prenatal DDE to midlife AD-relevant outcomes is absent.

**Objective:** To determine whether prenatal DDE exposure is associated with plasma AD biomarkers and cognitive performance in midlife, and whether APOE ε4 genotype modifies these associations.

**Design:** Observational cohort analysis nested within the Child Health and Development Studies (CHDS), a population-based birth cohort.

**Setting:** CHDS enrolled pregnant women between 1959-1967 in the San Francisco Bay Area.

**Participants:** Among 367 eligible adult offspring who participated in a follow-up study (2010-2013) at mean age 49.3 years, 179 with available prenatal DDE measurements were included.

**Main Outcomes and Measures:** Primary outcomes were prenatal DDE levels from maternal serum, plasma Aβ42/40 ratio and Digit Symbol Substitution Test (DSST) performance and APOE genotype. Secondary outcomes included plasma pTau217, GFAP, NfL and measures of verbal fluency (VF) and the Wechsler Test of Adult Reading (WTAR).

**Results:** Among 179 participants (56% female; 26% APOE ε4 carriers), mean prenatal DDE was 47.4 (25.4) ng/mL. Higher prenatal DDE was associated with lower DSST scores (β=−0.021; 95% CI, −0.041 to −0.001; P=0.039) and lower plasma Aβ42/40 ratio (β=−0.079; 95% CI, −0.133 to −0.024; P=0.005) per ng/mL DDE, adjusting for sex, race, education, and APOE ε4 status. Associations were strongest among APOE ε4 non-carriers for DSST (β=−0.033; 95% CI, −0.050 to −0.016; P=0.001) and Aβ42/40 ratio (β=−0.101; 95% CI, −0.161 to −0.040; P=0.001). No significant associations were observed for pTau217, GFAP, NfL, VF or WTAR.

**Conclusions and Relevance:** In this prospective birth cohort study, prenatal exposure to a persistent environmental toxicant was associated with lower plasma Aβ42/40 ratio and worse cognitive performance in midlife, consistent with DDE accelerating the preclinical trajectory of AD-related biological changes decades before symptom onset. These findings support a life-course framework for AD risk and identify prenatal DDE as a potentially modifiable determinant of early AD-related pathology amenable to prevention.

**Key Points:** *Question:* Is prenatal exposure to dichlorodiphenyldichloroethylene (DDE), primary persistent metabolite of DDT, associated with plasma Alzheimer disease biomarkers and cognitive performance in early midlife offspring?

*Findings:* In 179 participants from a prospective birth cohort followed for 50 years, higher prenatal DDE was associated with lower plasma Aβ42/40 and worse cognitive performance at mean age 49.3 years. Associations were substantially stronger among APOE ε4 non-carriers.

*Meaning:* Prenatal exposure to a ubiquitous environmental toxicant is associated with amyloid-related biomarker and cognitive changes in early midlife, consistent with an accelerated preclinical Alzheimer disease trajectory and supporting fetal environmental exposures as modifiable determinants of long-term AD risk.

## Introduction

Alzheimer disease (AD) is the leading cause of dementia worldwide and represents a growing public health challenge, impacting nearly 7 million individuals in the United States alone and 55 million globally.^1^ Increasing evidence indicates that AD develops over decades, with pathophysiologic changes, including amyloid-beta (Aβ) accumulation, beginning long before clinical symptoms emerge. Identifying determinants of early disease risk is therefore critical for prevention and early intervention.

The apolipoprotein E ε4 allele is the strongest genetic risk factor for Alzheimer’s disease, yet the majority of ε4 carriers never develop dementia, and heritable factors explain less than half of disease risk. What accounts for the remainder is unknown, but increasingly points to environmental exposures acting across the lifespan, alone and in interaction with genetic susceptibility, as modifiable determinants of who develops Alzheimer’s disease and when.^2,3^

Epidemiological evidence demonstrates that persistent organic pollutants such as the pesticide dichlorodiphenyltrichloroethane (DDT) and its primary metabolite dichlorodiphenyldichloroethylene (*p,p*_′_-DDE, hereafter DDE) are elevated in the brain and serum of AD patients compared to controls.^4–8^ Experimental studies demonstrate that DDT and DDE can influence amyloid-related pathways, including increased amyloid precursor protein and Aβ levels, providing biological plausibility for their role in AD pathogenesis.^4,9^ These observations raise the possibility that environmental exposures may contribute to AD risk independently or in conjunction with established genetic susceptibility. However, most studies of environmental contributions to AD risk have focused on exposure measurements obtained near the time of outcome assessment, limiting insight into whether earlier-life exposures contribute to disease development over time.

To address this gap, we examined whether prenatal exposure to DDE, measured in maternal serum during offspring gestation, was associated with subsequent midlife cognitive performance and plasma AD-related biomarkers. We also evaluated whether these associations differed by APOE ε4 status.

## Methods

### Study population

The Child Health and Development Studies (CHDS; chdstudies.org) enrolled nearly all pregnant mothers seeking obstetric care at the Kaiser Foundation Health Plan in Oakland, California, and the surrounding East Bay, from 1959 to 1967 (19,044 live births). This study is based on the adult offspring born into the CHDS from 1959 to 1967 who participated in a follow-up CHDS Disparities study (DISPAR) from 2010 to 2013 in early midlife (ages 45-52 years; N=605), which included an in-person visit and biospecimen collection.^10^ Plasma from participants who provided blood and consent for DNA analysis and inclusion in future studies (n=367) was used to determine AD/ADRD biomarkers. Of those, a subset of n=179 had available prenatal DDT and DDE measurements **(eFigure 1 in Supplement 1)**.^11^ The present study was reviewed and approved by the Institutional Review Board of the Public Health Institute (IRB# I22-013).

**Figure 1:**
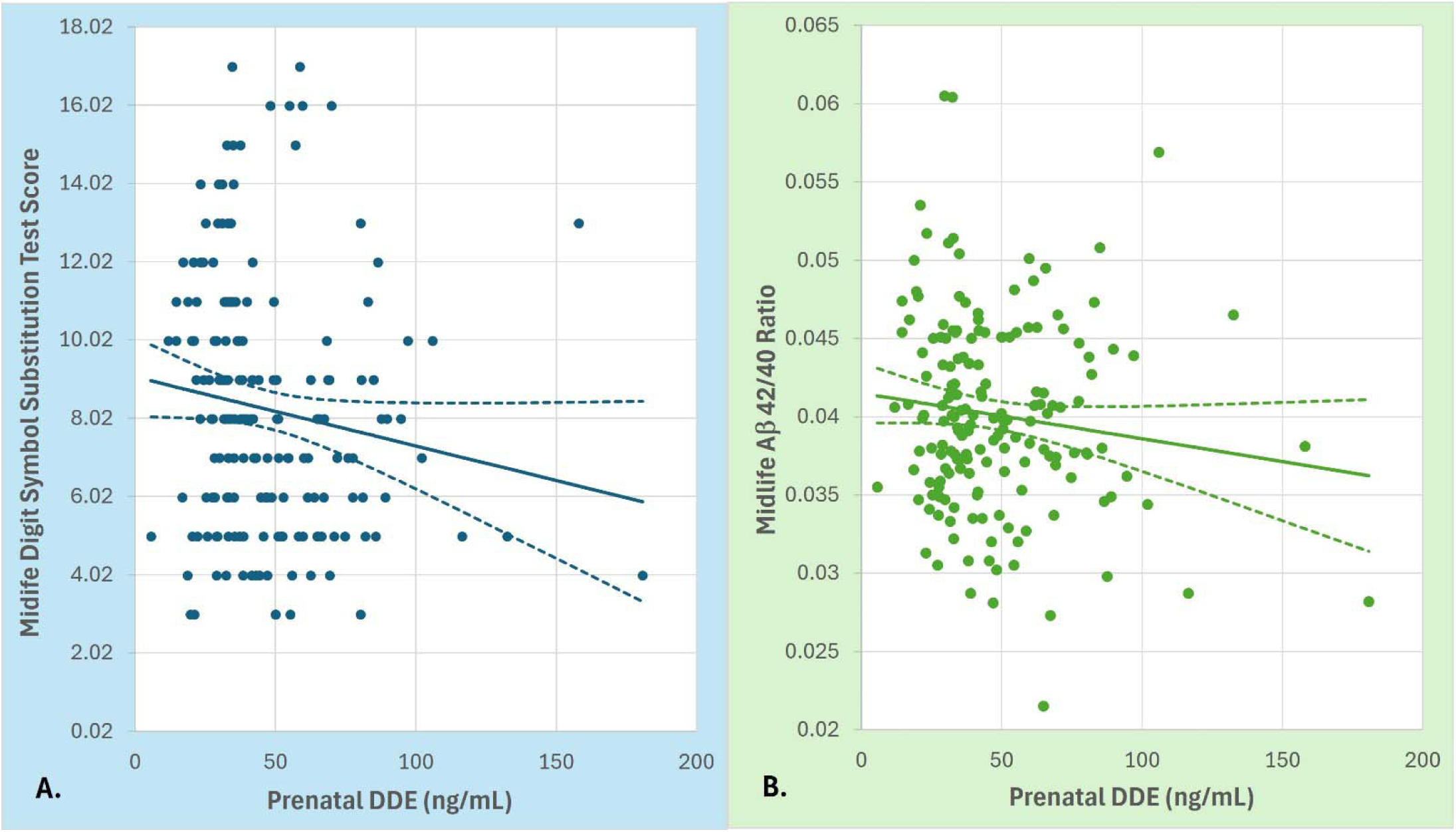
Scatter plots of (A) Digit Symbol Substitution Test score in mid-life and (B) Aβ 42/40 ratio in mid-life by level of prenatal DDE in the CHDS DISPAR Alzheimer follow-up study subset with available prenatal DDT measures, n=179.

### Assessment of DDT and DDE levels

Levels of maternal DDT congeners (DDT and DDE) available for the convenience sub-sample were assayed as previously described^11–13^ from frozen serum samples, drawn during pregnancy: 1% from the 1^st^ trimester, 2% from the 2^nd^ trimester, 4% from the 3^rd^ trimester, and 93% from the early postpartum, typically 1-3 days after birth. Previous evidence demonstrates measurement reliability over time^11^ and correspondence in levels across gestation^14^, supporting that the timing of specimen collection during the perinatal period is representative of pregnancy exposures.

### Cognitive Measurements

Cognitive tests were administered during home visits. For the Digit Symbol Substitution Test (DSST)^16^, we used standardized scores (mean = 10; standard deviation = 3) calculated from the count of numbers correctly translated to symbols.^17^ Test scores for the Verbal Fluency test (VF)^15^ and the Wechsler Test of Adult Reading (WTAR) were based on the number correct which was used for analysis.^16^ Administration of VF and WTAR was recorded and scored independently by two research staff, with excellent reliability (intraclass correlation = 0.989).^17^

### Plasma Biomarker Assays

Plasma Aβ40, Aβ42, total tau, phosphorylated tau 217 (p-Tau217_ALZpath_), glial fibrillary acidic protein (GFAP) and neurofilament light (NfL) levels were quantified on the Quanterix SR-X ultra-sensitive biomarker detection system using single-molecule array (Simoa) technology. Plasma p-Tau217 concentration was also quantified using the ultrasensitive S-plex Human Tau (p-Tau217_Lilly_) kit on the Meso Scale Discovery platform. QA/QC measures are provided in **eTable 1 in Supplement 1**.

### APOE Genotyping

APOE genotype was determined by polymerase chain reaction based allelic discrimination of single-nucleotide polymorphisms rs429358 and rs7412 using TaqPath™ ProAmp Master Mix (Thermo Fisher Scientific). Genotypes were assigned based on allele-specific amplification. A random 10% subset of samples was validated using modified Sanger sequencing.

### Statistical Analyses

Statistical analyses were performed using SAS version 9.4 (SAS Institute Inc). DDT and DDE were characterized as continuous variables, natural log-transformed variables, and as tertiles using 2 dummy variables representing tertile 2 and tertile 3, versus tertile 1 (the lowest) as the reference category.

Cognitive function test scores were classified as continuous variables and as natural log-transformed variables. AD/ADRD biomarkers were similarly classified as continuous variables (pg/mL or ratio units) and as natural log-transformed variables. Both DSST and the Aβ 42/40 ratio were additionally assessed as dichotomous outcomes (lower 40% vs upper 60%).

We used linear and logistic models to estimate associations, 95% confidence limits, and tests of statistical significance (p-value <0.05). Generalized linear models with robust variance estimation were used to account for correlation between sibling pairs in the study (N = 8). Fully adjusted models included potential confounders based on a directed acyclic graph (DAG); variables included were age, sex at birth, race (Black, non-Black), completed years of education and DDT concentration (ng/mL).

To evaluate effect modification by APOE ε4 status, interaction terms between DDE and APOE ε4 carrier status were included in fully adjusted models for DSST and Aβ42/40, followed by stratified analyses in APOE ε4 carriers and non-carriers.

### Additional Analyses

Sensitivity analyses were conducted to determine the impact of excluding siblings and the timing of pregnancy blood draw (see eSupplemental Methods).

## Results

Participant characteristics are shown in **Table 1**. Details for assay performance and the distribution of AD-related plasma biomarkers across the study population are shown in **eTables 1 and 2 in Supplement 1**.

**Table 1.** Distribution of study sample characteristics.

Main Tables
| Table 1. Distribution of study sample characteristics |  |  |  |  |  |
| --- | --- | --- | --- | --- | --- |
| Study Sample Characteristics | Mean (SD)<br>or N (%) | Range<br>(Min-Max) | Quartiles |  |  |
|  |  |  | 25<sup>th</sup> | 50<sup>th</sup> | 75<sup>th</sup> |
| Biological Sex at Birth |  |  |  |  |  |
| Female | 100 (56%) | ----- | ----- | ----- | ----- |
| Male | 79 (44%) | ----- | ----- | ----- | ----- |
| Race |  |  |  |  |  |
| Black | 47 (26%) | ----- | ----- | ----- | ----- |
| Non-Black | 132 (74%) | ----- | ----- | ----- | ----- |
| APOE Genotype |  |  |  |  |  |
| APOE ε4 carriers (+) | 47 (26%) | ----- | ----- | ----- | ----- |
| APOE ε4 non-carriers (-) | 132 (74%) | ----- | ----- | ----- | ----- |
| Other Mid-life Characteristics |  |  |  |  |  |
| Year of Birth | 1961.4 (1.0) | 1960 – 1966 | 1961 | 1961 | 1962 |
| Age at Mid-Life Study (y) | 49.3 (1.0) | 45.7 – 51.6 | 48.7 | 49.4 | 50.1 |
| Completed Education (y) | 15.1 (2.6) | 11 – 21 | 13 | 16 | 16 |
| Prenatal DDE and DDT levels |  |  |  |  |  |
| *p,p'*-DDE (ng/mL) | 47.4 (25.4) | 5.7 – 180.8 | 31.2 | 39.2 | 60 |
| *p,p'*-DDT (ng/mL) | 14.9 (10.1) | 0.6 – 52.5 | 8.3 | 11.4 | 18.3 |
| Mid-life Plasma Biomarkers |  |  |  |  |  |
| Aβ40 (pg/ml) | 174.7 (51.2) | 39.6 – 459.8 | 142.8 | 170.7 | 192.7 |
| Aβ42 (pg/ml) | 6.8 (1.9) | 2.1 – 15.5 | 5.6 | 6.7 | 7.7 |
| Total Tau (pg/ml) | 2.4 (1.2) | 0.2 – 10.8 | 1.7 | 2.1 | 2.8 |
| p-Tau217<sub>ALZpath</sub> (pg/ml) | 0.19 (0.10) | 0.01 – 0.81 | 0.13 | 0.17 | 0.22 |
| p-Tau217<sub>Lilly</sub> (pg/ml) | 3.1 (2.2) | 0.02 – 9.76 | 1.5 | 2.7 | 4.4 |
| GFAP (pg/ml) | 68.4 (39.2) | 18.4 – 363.5 | 44.2 | 61.9 | 80.6 |
| NfL (pg/mL) | 7.5 (3.0) | 2.7 – 19.1 | 5.4 | 7.2 | 8.9 |
| Aβ42/40 ratio | 0.040 (0.008) | 0.009 – 0.086 | 0.036 | 0.040 | 0.045 |
| p-Tau217<sub>ALZpath</sub> / Aβ42 ratio | 0.03 (0.02) | 0.003 – 0.203 | 0.019 | 0.026 | 0.035 |
| p-Tau217<sub>Lilly</sub> / Aβ42 ratio | 0.5 (0.5) | 0.00 – 4.06 | 0.2 | 0.4 | 0.7 |
| Cognitive Tests |  |  |  |  |  |
| Wechsler Digit Symbol Substitution Test (DSST) | 8.2 (3.3) | 3 – 17 | 6 | 8 | 10 |
| Wechsler test of Adult Reading (WTAR) | 101.7 (14.8) | 59 – 123 | 94.3 | 105 | 113 |
| Verbal Fluency (VF) | 23.6 (6.3) | 7.0 – 41.0 | 19 | 24 | 28 |
| Abbreviations: APOE, Apolipoprotein E; Aβ - Amyloid Beta peptide;<br>p-Tau217 - phosphorylated tau protein at the threonine 217 position; GFAP - Glial Fibrillary Acidic Protein;<br>NfL - Neurofilament Light chain protein; BD-Tau - Brain Derived Tau protein. |  |  |  |  |  |

### Associations between prenatal DDE and midlife cognitive function

Prenatal DDE concentrations were associated with lower midlife cognitive performance on the DSST **(Figure 1A)**. In continuous models adjusted for sex and race, higher DDE levels were associated with lower DSST scores (β = −0.022; 95% CI, −0.040 to −0.004; P = 0.018), with similar results observed after full adjustment including education (β = −0.021; 95% CI, −0.041 to −0.001; P = 0.039) **(Table 2)**.

**Table 2.** Associations between prenatal DDE and mid-life cognition and Aβ42/40.

| Outcomes and Predictors<sup>1</sup> | Sex and Race Adjusted |  |  |  | Fully Adjusted |  |  |  |
| --- | --- | --- | --- | --- | --- | --- | --- | --- |
|  | Estimate | 95% Confidence Limit |  | P-value | Estimate | 95% Confidence Limit |  | P-value |
|  |  | Lower | Upper |  |  | Lower | Upper |  |
| 1. Outcome: continuous Digit Symbol Substitution Test (DSST) per 1 unit<sup>2</sup> |  |  |  |  |  |  |  |  |
| Continuous DDE (ng/mL) | -0.022 | -0.040 | -0.004 | 0.018 | -0.021 | -0.041 | -0.001 | 0.039 |
| 2. Outcome: continuous Aβ42/40 Ratio per 1,000 units<sup>3</sup> |  |  |  |  |  |  |  |  |
| Continuous DDE (ng/mL) | -0.078 | -0.133 | -0.023 | 0.005 | -0.079 | -0.133 | -0.024 | 0.005 |

<sup>1</sup>Associations were estimated using generalized linear models and binomial logistic models as appropriate to the parameterization of the outcomes and adjusted for sibling clusters. For logistic models, estimates, rather than exponentiated odds ratios are reported to provide consistency with linear models.
<sup>2</sup>Sex and race-adjusted models included F1 biological sex at birth (male vs female) and race (Black vs non-Black). Fully adjusted model added completed education (continuous years).
<sup>3</sup>Sex and race-adjusted models included F1 biological sex at birth (male vs female), race (Black vs non-Black) and *p,p'*-DDT (continuous ng/mL). Fully adjusted model added completed education (continuous years).

In categorical analyses, participants in DDE tertile 2 (β = −0.876; 95% CI, −1.6995 to −0.0525; P = 0.04) and tertile 3 (β = −1.3268; 95% CI, −2.31 to −0.3436; P = 0.008) had lower DSST scores than those in tertile 1. These associations persisted after full adjustment, tertile 2 (β = −1.129; 95% CI, −2.048 to −0.209; P = 0.016) and tertile 3 (β = −1.596; 95% CI, −2.657 to −0.5347; P = 0.003) **(eTable 3 in Supplement 1)**. A significant trend was observed across DDE tertiles (P for trend < .01).

No significant associations were observed between prenatal DDE levels and scores on the WTAR or VF test **(eTable 4 in Supplement 1)**.

### Association of prenatal DDE with midlife A**β**42/40 ratio

Higher prenatal DDE concentrations were associated with lower plasma Aβ42/40 **(Figure 1B)**, but not other plasma AD biomarkers **(eTable 5 in Supplement 1).** In continuous models scaled per 1,000 units, higher DDE levels were associated with lower Aβ42/40 ratios in sex and race-adjusted models (β = −0.078; 95% CI, −0.133 to −0.023; P = 0.005), with similar results observed after full adjustment including education (β = −0.079; 95% CI, −0.133 to −0.024; P = 0.005) **(Table 2)**.

When modeled categorically, higher DDE levels were associated with lower plasma Aβ42/40 ratio. In sex and race-adjusted models, compared to tertile 1 (reference), tertile 2 (β = −0.882; 95% CI, −1.763 to 0.001; P = 0.05) and tertile 3 (β = −1.018; 95% CI, −2.021 to −0.014; P = 0.047) were associated with lower Aβ42/40. Adjusting for education, estimates remained directionally similar but were slightly attenuated, with borderline statistical significance for tertile 2 (β = −0.855; 95% CI, −1.730 to 0.021; P = 0.056) and tertile 3 (β = −1.000; 95% CI, −2.013 to 0.014; P = 0.053) **(eTable 3 in Supplement 1)**. A significant trend was observed across DDE tertiles (P for trend = 0.05).

Results were robust to different parameterizations of predictor and outcome. Log-transformed DDE produced similar results with both DSST and Aβ42/40 suggesting there was no undue influence from outlier DDE values **(eTable 3 in Supplement 1)**. The step-down gradient in DSST scores and Aβ42/40 levels with increasing DDE tertiles suggests a dose-response effect for both outcomes, that is also consistent with the linear models **(eTable 3 in Supplement 1)**. Exclusion of siblings and adjustment for DDT levels measured within the 1st-3rd trimesters, as well as adjustment for the timing of maternal pregnancy blood draw, did not alter DDE associations with either DSST or Aβ42/40 **(eTable 3 in Supplement 1)**.

### APOE status modifies the association of prenatal DDE with DSST and plasma A**β**42/40

Participants were grouped as APOE ε4 non-carriers (ε2/ε2, ε2/ε3, and ε3/ε3) or carriers (ε2/ε4, ε3/ε4, and ε4/ε4) **(eTable 6 in Supplement 1)**. Prenatal DDE levels did not differ by APOE ε4 carrier status **(eTable 7 in Supplement 1)**.

The association between prenatal DDE and DSST performance differed by APOE ε4 status (P_interaction_ = 0.038 for DDE X APOE ε4 interaction), with a negative association observed among non-carriers but not among carriers **(Figure 2A**; **Table 3)**. Among APOE ε4 non-carriers, higher prenatal DDE exposure was associated with progressively lower mean DSST scores (β = −0.033; 95% CI = −0.050 to −0.016; P = < 0.01), while DSST scores did not differ across DDE tertiles among ε4 carriers (β = 0.004; 95% CI = - 0.029 to 0.037; P = NS). The interaction model **(Table 3)** was used to estimate mean DSST levels by DDE tertile (at the median) for APOE ε4 non-carriers vs. carriers **(Figure 2A**). As depicted in Figure 2A higher DDE tertiles were associated with lower DSST levels in non-carriers but not in carriers. Stratified analyses confirmed these results **(Table 3)**.

**Figure 2.**
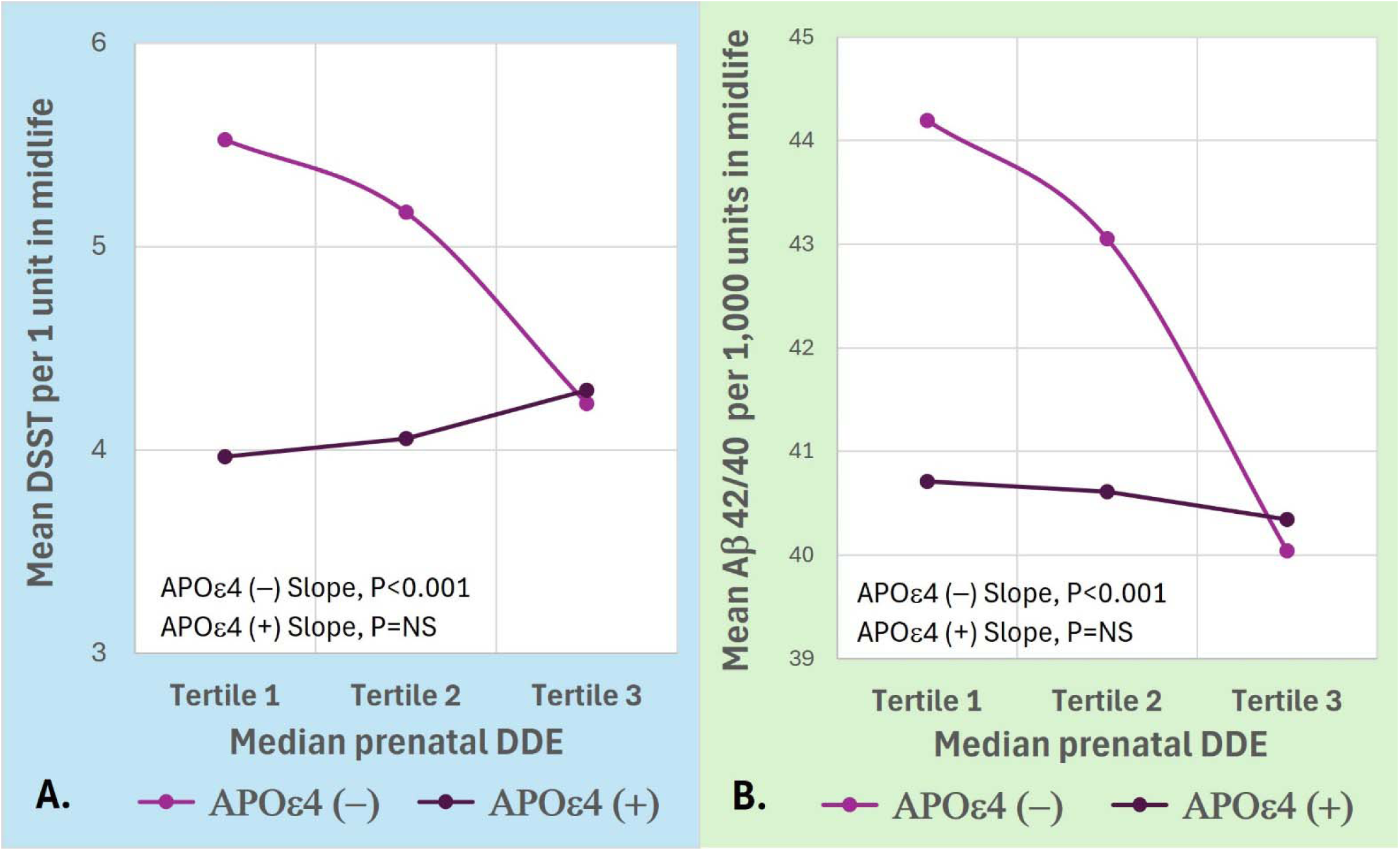
Estimated midlife DSST scores (A) and plasma Aβ42/40 ratios (B) across tertiles of prenatal DDE exposure, stratified by APOE ε4 carrier status (APOE ε4 non-carriers, N=132 and APOE ε4 carriers, N=47). Estimates were derived from generalized linear models adjusted for sibling clustering, sex, race, educational attainment, and, for Aβ42/40, prenatal DDT.

**Table 3.** APOE ε4 polymorphism modifies the associations of mid-life cognition and Aβ42/40 with DDE.

| Outcomes and Predictors<sup>1</sup> | Estimate | 95% Confidence Limit |  | P-value |
| --- | --- | --- | --- | --- |
|  |  | Lower | Upper |  |
| 1. Outcome: continuous Digit Symbol Substitution Test (DSST) per unit<sup>2</sup> |  |  |  |  |
| Independent Effects Model |  |  |  |  |
| Continuous DDE (ng/mL) | -0.020 | -0.041 | 0.001 | 0.058 |
| APOE ε4 allele positive vs negative | -0.677 | -1.617 | 0.264 | 0.159 |
| Interaction Model |  |  |  |  |
| Continuous DDE (ng/mL) | -0.033 | -0.049 | -0.016 | <.0001 |
| APOE ε4 allele positive (vs negative) | -2.699 | -4.893 | -0.505 | 0.016 |
| DDE (ng/mL) X APOE ε4 allele positive | 0.041 | 0.002 | 0.079 | 0.038 |
| DDE effect in Models Stratified by APOE ε4 Allele |  |  |  |  |
| APOE ε4 allele positive, n=47 | 0.004 | -0.029 | 0.037 | 0.821 |
| APOE ε4 allele negative, n=131 | -0.033 | -0.050 | -0.016 | 0.001 |
| 2. Outcome: continuous Aβ42/40 Ratio per 1,000 units<sup>3</sup> |  |  |  |  |
| Independent Effects Model |  |  |  |  |
| Continuous DDE (ng/mL) | -0.078 | -0.133 | -0.022 | 0.006 |
| APOE ε4 allele positive vs negative | -1.432 | -3.612 | 0.749 | 0.198 |
| Interaction Model |  |  |  |  |
| Continuous DDE (ng/mL) | -0.104 | -0.164 | -0.043 | 0.001 |
| APOE ε4 allele positive (vs negative) | -6.130 | -10.631 | -1.629 | 0.008 |
| DDE (ng/mL) X APOE ε4 allele positive | 0.095 | 0.007 | 0.182 | 0.034 |
| DDE effect in Models Stratified by APOE ε4 Allele |  |  |  |  |
| APOE ε4 allele positive, n=47 | -0.031 | -0.165 | 0.103 | 0.652 |
| APOE ε4 allele negative, n=132 | -0.101 | -0.161 | -0.040 | 0.001 |

<sup>1</sup>Associations were estimated using generalized linear models adjusted for sibling clusters.
<sup>2</sup>Models were adjusted for F1 biological sex at birth (male vs female), race (Black vs non-Black) and completed education (continuous years).
<sup>3</sup>Models were adjusted for F1 biological sex at birth (male vs female), race (Black vs non-Black), p,p'-DDT (continuous ng/mL) and completed education (continuous years).

The association between DDE and Aβ42/40 ratio (per 1,000 units) also differed by APOE ε4 status **(Figure 2B and Table 3**, P_interaction_ = 0.034). In APOE ε4 non-carriers, prenatal DDE exposure was associated with declining Aβ42/40 (β = −0.101; 95% CI = −0.161 to −0.040; P < 0.01). In contrast, Aβ42/40 ratios remained relatively stable across DDE tertiles among ε4 carriers (β= −0.031; 95% CI = - 0.165 to 0.103; P = NS). For Aβ42/40, the interaction model **(Table 3)** was used to estimate mean Aβ42/40 levels by DDE tertile for APOE ε4 non-carriers vs. carriers **(Figure 2B)**. As depicted in Figure 2B, higher DDE tertiles were associated with lower Aβ42/40 levels in non-carriers but not in carriers. Stratified analyses corroborated these results **(Table 3)**.

## Discussion

With the growing global burden of Alzheimer disease (AD), identifying early-life determinants of disease risk has become increasingly important. Most environmental epidemiologic studies of AD have relied on exposure measurements obtained in late life, limiting insight into whether earlier-life exposures contribute to disease development, which limits opportunities for early intervention. In this population-based birth cohort, higher prenatal exposure to dichlorodiphenyldichloroethylene (DDE) was associated with lower cognitive performance and reduced plasma Aβ42/40 ratio in offspring at midlife (mean age, 49.3 years). These results indicate that AD-related biological processes may be initiated during fetal development, shifting the temporal origin of disease risk decades earlier than currently recognized, providing an opportunity for precision prevention and risk stratification.

In this pregnancy cohort, maternal prenatal serum DDE concentrations were consistent with ubiquitous and high DDT use in the 1960’s, supporting the relevance of prenatal exposure to population health.^18^ Our findings apply to midlife AD-biomarkers, likely representing early manifestations along the Alzheimer disease continuum, and reflect prospective associations between early life exposures and midlife outcomes. Prior epidemiologic studies have found higher levels of serum or plasma DDE in those with AD and associations with poorer cognitive performance in older adults^4,19^; however, these investigations have largely relied on exposure measurements obtained in older age or after clinical onset. Experimental studies further demonstrate that DDT and DDE within the range of human exposures during the 1960s can increase amyloid precursor protein expression^4,9^ and promote amyloid deposition.^4^ In addition, prior work has demonstrated a strong correlation between serum and brain concentrations of DDE, supporting the biological relevance of circulating measures as a proxy for central nervous system exposure.^4^ Together, these data support a biologically plausible pathway linking DDE exposure to increased amyloid precursor protein expression and Aβ levels, providing mechanistic context for the observed human associations and supporting the conclusion that DDE is neurotoxic rather than simply a proxy for prior DDT exposure.

The observed association between prenatal DDE exposure and lower Digit Symbol Substitution Test (DSST) performance in midlife is consistent with prior reports linking DDT/DDE exposure to cognitive dysfunction. However, previous studies measured exposure at older ages often in occupationally exposed populations^4,5,19^ In contrast, this study investigated prenatal exposure in relation to midlife outcomes in a community-based cohort. Processing speed and executive function, as assessed by the DSST, are sensitive to early disruption of large-scale brain networks and may represent among the earliest detectable functional changes along the preclinical or prodromal stages of Alzheimer disease.^20–24^ The concurrent association with a lower plasma Aβ42/40 ratio in this cohort suggests early disruption of pathophysiologic processes in association with prenatal DDE exposures rather than age-related cognitive decline alone.

The plasma Aβ42/40 ratio represents one of the earliest detectable biological alterations along the AD continuum, often in the preclinical phase, preceding clinical symptoms by almost decades.^24–26^ The association between higher prenatal DDE exposure and lower Aβ42/40 ratio in midlife suggests that early-life environmental exposures may be associated with amyloid-related processes decades before clinical onset. The specificity of these findings to Aβ42/40, in the absence of downstream neurodegenerative markers, is consistent with established temporal models of Alzheimer disease in which amyloid alterations precede tau-related injury and neuronal loss.^27,28^ These results are consistent with early preclinical stages of Alzheimer disease and support the interpretation that prenatal exposures may influence the timing of initial amyloid-related changes.^26,29–31^ In addition, the integration of objective exposure measures with plasma biomarkers and standardized cognitive testing in a community-based population further strengthens the relevance of these findings to population health.

Associations between prenatal DDE exposure and both cognitive performance and Aβ42/40 ratio were more pronounced among individuals without the APOE ε4 allele. These findings support the possibility that environmental exposures may contribute to AD risk through pathways partially independent of APOE-mediated susceptibility, although this interpretation requires confirmation in larger cohorts. One possible explanation is that baseline differences in plasma Aβ42/40 ratio and cognitive performance among ε4 carriers may reduce the dynamic range for detecting additional exposure-related effects at this stage of life. This explanation is supported by our observation that plasma Aβ42/40 ratio levels and cognitive performance are much lower for ε4 carriers than non-carriers at low levels of DDE but converge at high DDE levels. Alternatively, environmental exposures may exert greater relative influence among individuals without strong genetic risk. These findings highlight the importance of considering gene-environment interactions across the life course but require confirmation in larger cohorts.

### Limitations

Several limitations should also be considered. Although the prospective design establishes temporal separation between exposure and outcome and assesses exposure during development, a sensitive window for brain development, these findings should be interpreted as associations and do not establish causality. However, the follow-up spanning more than five decades provides a rare opportunity to examine associations between early-life exposures and midlife cognition and Alzheimer’s biomarkers. Residual confounding by socioeconomic factors, maternal health, diet, or co-exposures to other pollutants cannot be excluded, as DDE may serve as a marker of other exposures. However, the DDE associations are consistent with experimental studies showing significant impact on amyloid levels at concentrations similar to those observed here. Although the sample size was modest, the consistency of associations across continuous and categorical models, along with concordant cognitive and biomarker findings, supports the robustness of the observed relationships. While the number of APOE _ε_4 carriers was small, we still found a significant interaction, suggesting that limited statistical power was not an issue. Plasma biomarkers and cognitive outcomes were assessed at a single midlife time point, precluding evaluation of longitudinal trajectories. The offspring cohort are now entering their early to late 60s, a time when the risk for cognitive decline and dementia begins to rapidly increase, providing a unique opportunity to examine the long-term consequences of prenatal exposure across the life course, which we plan to follow up. The absence of confirmatory cerebrospinal fluid or neuroimaging measures is also a limitation. Finally, we did not examine contemporary midlife exposures that might act in synergy with prenatal DDE.

## Conclusions

Taken together, these findings provide human evidence that prenatal exposure to a persistent environmental toxicant is associated with cognitive performance and amyloid-related biomarkers measurable nearly five decades later, supporting a life-course model of AD risk. By linking prenatal exposure to early midlife signatures of preclinical disease, these results indicate that environmental exposures during fetal development may shift the timing of early AD-related biological processes decades before clinical onset. Although effect sizes were modest, they are consistent with subtle biologic changes that precede clinical AD by decades and may accumulate over time to influence later risk. The observation that associations were more pronounced among individuals without APOE ε4 further underscores the potential for environmental exposures to contribute to disease pathways beyond established genetic susceptibility. If confirmed, these findings support a model in which modifiable environmental exposures during critical developmental windows contribute to the earliest stages of AD biology, with implications for prevention strategies beginning in the prenatal period.

## Author Contributions

Study concept and design: Richardson, Cohn, Jones, and Factor-Litvak

Acquisition, analysis, or interpretation of data: All authors

Drafting of the manuscript: Mhatre-Winters, Cirillo, and Richardson

Critical review of the manuscript for important intellectual content: All authors

Statistical analysis: Cirillo, Mhatre-Winters, and Factor-Litvak

Obtained funding: Richardson, Cohn, Jones, Link, and Factor-Litvak

Administrative, technical, or material support: Krigbaum, Han, and Zimmermann

Supervision: Richardson, Cohn, Jones, and Factor-Litvak

## Data Sharing Statement

De-identified (anonymized) data are available upon request from Barbara A. Cohn, PhD, Director of the Child Health and Development Studies. Requests will be reviewed by Dr. Cohn, research staff, and the Institutional Review Board at the Public Health Institute. Approval of requests for de-identified (anonymized) data requires execution of a data use agreement.

## Additional Contributions

We thank the CHDS participants, especially those who co-enrolled in the DISPAR ancillary study.

