## Supplement 1 for "Prenatal Pesticide Exposure and Early Alzheimer Disease-Related Biomarker and Cognitive Changes in Midlife"

eFigure 1. Flow Chart of Study Participants

eTable 2. Distributions of plasma biomarkers in mid-life

eTable 3. Results of sensitivity analyses on associations of midlife Digit Symbol Substitution Test (DSST) and plasma Aβ42/40 with prenatal DDE

eTable 4. Associations between prenatal DDE and mid-life cognitive function as measured by the Wechsler Test of Adult Reading (WTAR) and the Verbal Fluency (VF) test

eTable 5. Associations of prenatal DDE with mid-life plasma biomarkers, n=179

eTable 6. Distribution of APOE genotype, n=179

eTable 7. Mean prenatal DDE levels stratified by APOE ε4 status

eMethods

### eFigure 1. Flow Chart of Study Participants

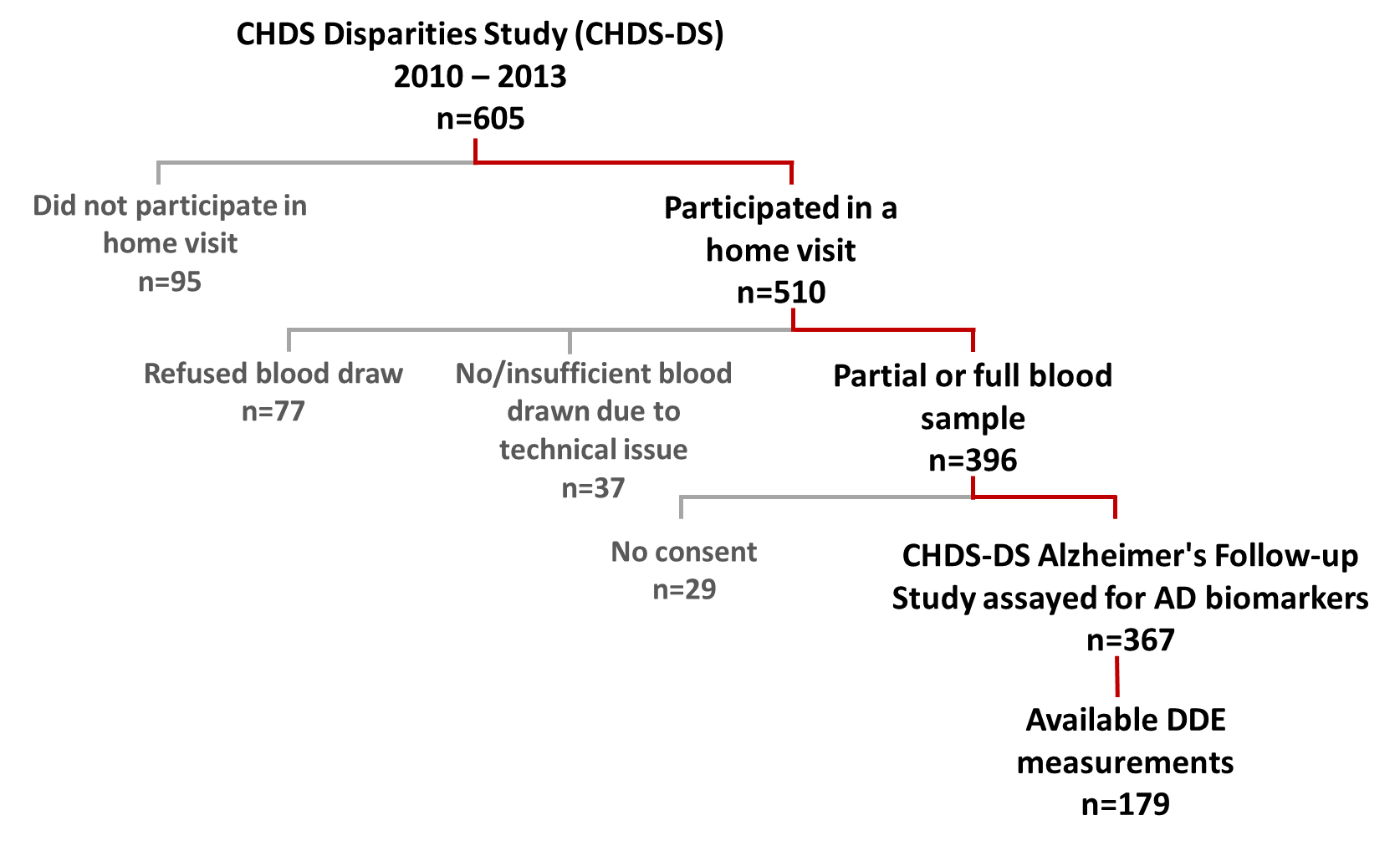

**eFigure 1.** Flow chart illustrating the derivation of the CHDS DISPAR Alzheimer's Follow-up Study (n=367)

### eTable 1. Assay analytics and LODs for plasma biomarkers measured in midlife

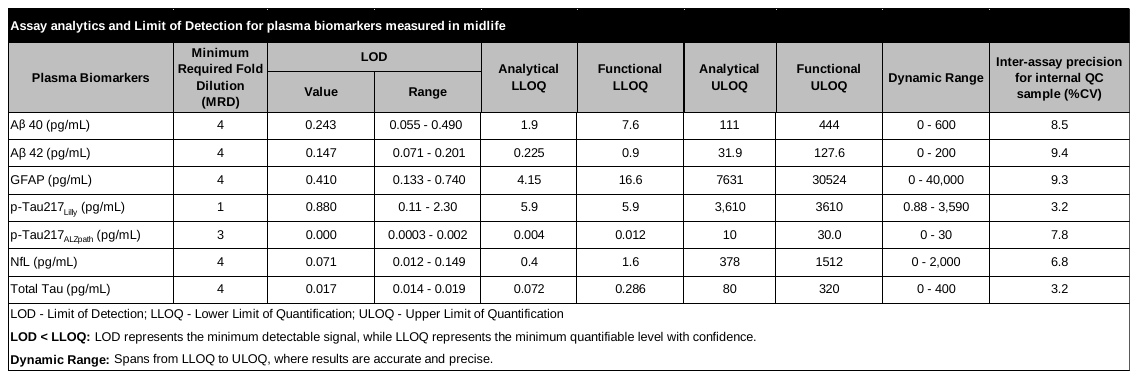

### eTable 2. Distributions of plasma biomarkers in mid-life

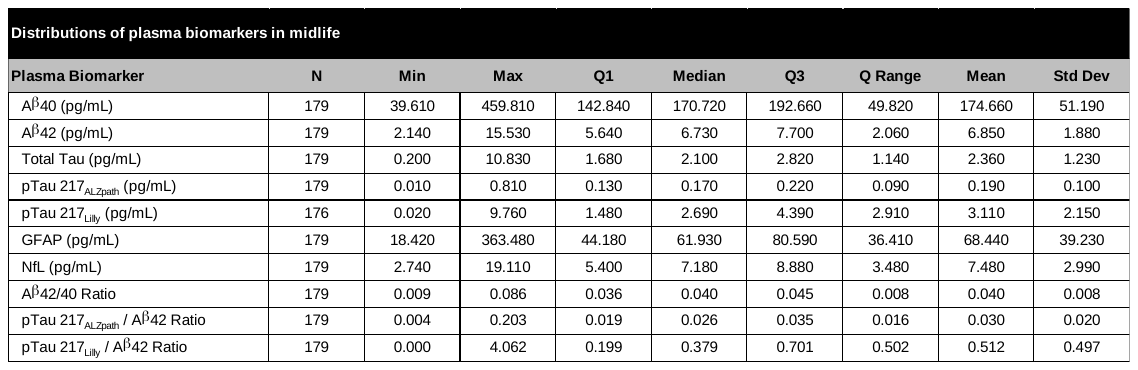

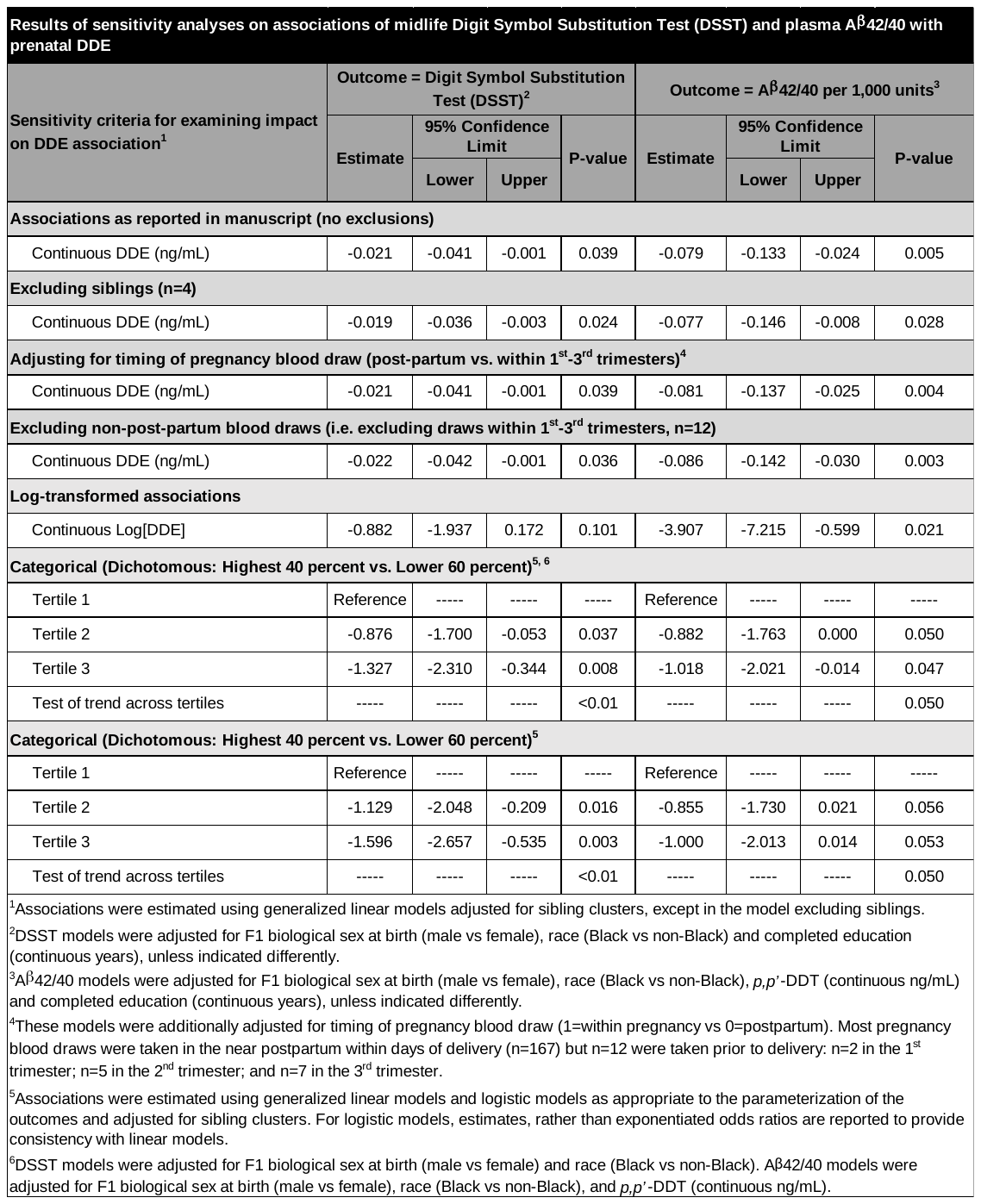

### eTable 3. Results of sensitivity analyses on associations of midlife Digit Symbol Substitution Test (DSST) and plasma Aβ42/40 with prenatal DDE

### eTable 4. Associations between prenatal DDE and mid-life cognitive function as measured by the Wechsler Test of Adult Reading (WTAR) and the Verbal Fluency (VF) test

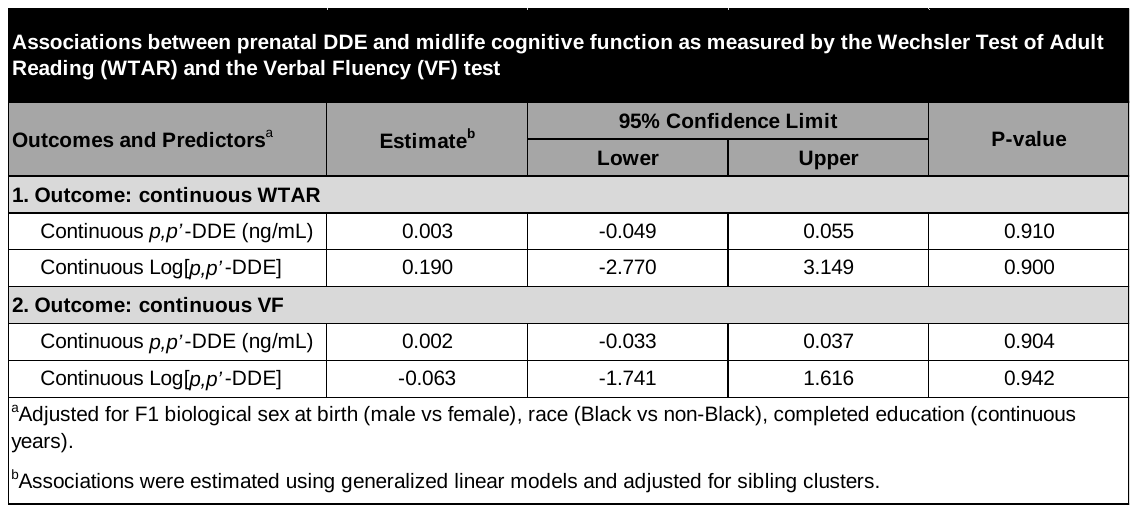

### eTable 5. Associations of prenatal DDE with mid-life plasma biomarkers, n=179

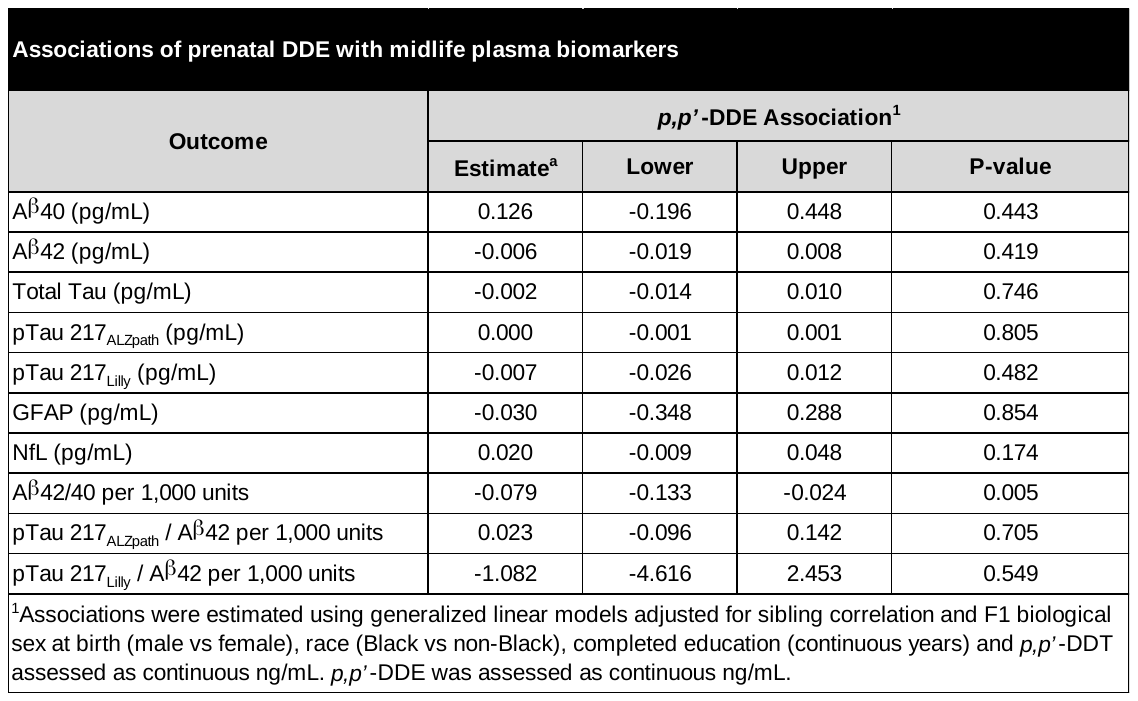

### eTable 6. Distribution of APOE genotype, n=179

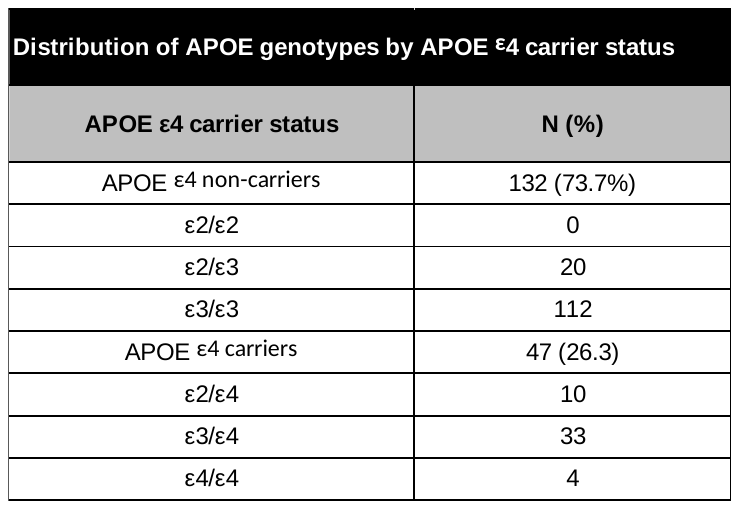

### eTable 7. Mean prenatal DDE levels stratified by APOE ε4 status

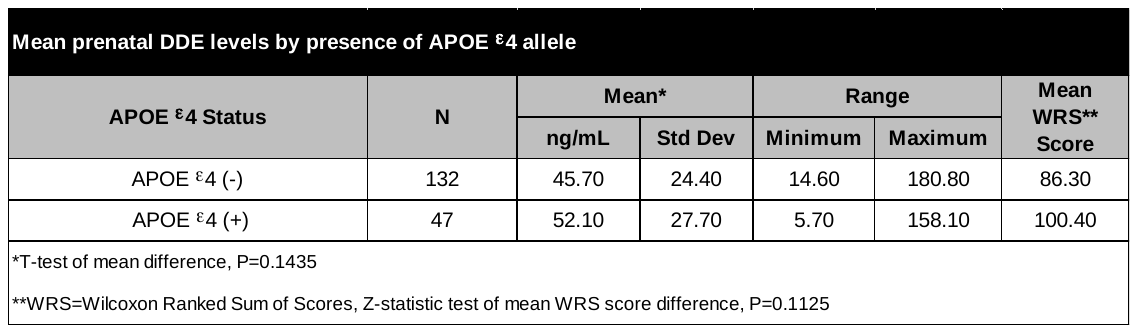

### eMethods

#### 1. Study population

The Child Health and Development Studies (CHDS; chdstudies.org) enrolled nearly all pregnant mothers seeking obstetric care at the Kaiser Foundation Health Plan in Oakland, California, and the surrounding East Bay, from 1959 to 1967 (19,044 live births).^1^ This study is based on the adult offspring born into the CHDS from 1959 to 1967, who participated in a series of childhood studies from ages 5 through adolescence^2^ and participated in a follow-up CHDS Disparities study (DISPAR) in 2010 at midlife adulthood (ages 45-52 years), which included an in-person visit and biospecimen collection.^3^

Demographic and health-related behavior of the parents were recorded in an in-person interview with the mother. Adult offspring recruited to the CHDS DISPAR study (n=605) completed midlife assessments of socioeconomic status, mental health, physical activity, and health behaviors, which were documented via telephone interviews. Biospecimens and anthropometric, blood pressure, spirometric, and cognitive function measurements were collected during an in-person visit.^3^ The current study uses the plasma collected from CHDS DISPAR participants who gave blood and consent (n=367) to measure levels of midlife prodromal AD biomarkers. Of those with measured AD biomarkers, a subset of n=179, with available prenatal *p,p’*-DDT and *p,p’*-DDE levels, were derived from the existing CHDS assay library of organochlorine measures accumulated over the course of multiple studies.^4^

This study was conducted in accordance with the ethical principles outlined in the Declaration of Helsinki. The present study was reviewed and approved by the Institutional Review Board of the Public Health Institute (IRB# I22-013). At enrollment CHDS mothers (F0) gave informed oral consent, as was customary in the 1960s, for themselves and their children (F1). F1 who participated in the DISPAR Study from 2010 to 2013 gave full informed verbal consent before completing the study surveys and written consent before participating in the home visits, including consent for use of DNA and inclusion in future studies.

#### 2. Covariates

Covariates included F1 biological sex assigned at birth (male or female), race (black or non-black), years of education, and prenatal *p,p′*-DDT. We used the most parsimonious, best-fitting models to estimate DDE associations with DSST and Aβ42/40, minimally including only covariates having known, *a priori* associations with these outcomes, i.e., age, race, APOE ε4, and education. DDT was also included in the Aβ42/40 models as a confounder of the DDE association but was excluded from the DSST models because of high collinearity with DDE.

#### 3. Cognitive Measurements

Test scores for the Digit Symbol Substitution Test (DSST), the Wechsler Test of Adult Reading (WTAR), and the Verbal Fluency (VF) test, administered in-person at ages 45-52 years, were used to investigate midlife cognitive function. The DSST is a timed test requiring rapid processing of unique symbols and their corresponding numbers or letters, and is sensitive to general brain dysfunction as well as normal age-related decline in function.^5^ It is a measure of fluid intelligence since it relies on the capacity to process complex information involved in reasoning and problem-solving tasks. The VF test is a second measure of fluid intelligence that requires participants to name as many animals as possible within 1 minute.^6^ The WTAR test requires participants to read a list of phonetically incorrect words and is used to assess crystallized intelligence.^7^

#### 4. Plasma biomarker assays

Frozen plasma EDTA samples were thawed and centrifuged at 10,000 g for 10 min at 4°C. Supernatants were aliquoted and frozen at -80°C. Assays were run blinded, and each sample was measured in duplicates. Measurements were performed using reagents from the same lot for each assay kit.

Plasma glial fibrillary acidic protein (GFAP) and neurofilament light (NfL) concentrations were measured with a Neurology 2-Plex B assay kit (Quanterix; item # 103520). Aβ40, Aβ42, and total tau concentrations were measured using a Neurology 3-Plex A assay kit (Quanterix; item # 101995). Phosphorylated tau 217 (p-Tau217_ALZpath_) levels were quantified using the ALZpath p-Tau217 Advantage PLUS Kit (Quanterix; item # 104570). All assays were a 2-step digital immunoassay performed according to the manufacturer’s instructions on a Quanterix SR-X ultra-sensitive biomarker detection system using single-molecule array (Simoa) technology.

Plasma p-Tau217 concentration was also quantified using the ultrasensitive S-plex Human Tau (p-Tau217_Lilly_) kit on the Meso Scale Discovery platform (MSD; catalog # K151APFS-2). Plates were read by electrochemiluminescence on the MSD SQ120 instrument.

The concentrations of all analytes were measured using an 8 or 9-point calibration curve. A 1/y^2^ weighted 4PL-fit algorithm was used to calculate sample concentrations for all analytes in pg/mL. The precision of analyte measurement for each assay on both platforms was determined by calculating the coefficient of variation (%CV) between technical replicates on one plate (intra-assay precision). An in-house pooled plasma sample was added to each plate as quality control to measure %CV across plates (inter-assay precision).

#### 5. DNA extraction and APOE Genotyping

Genomic DNA was extracted from 200 µL buffy coat samples using an automated protocol on the KingFisher Flex system (Thermo Fisher Scientific). APOE genotype was determined by polymerase chain reaction (PCR) based allelic discrimination of single-nucleotide polymorphisms rs429358 and rs7412 using TaqPath™ ProAmp Master Mix (Thermo Fisher Scientific) in 5 µL reactions on a QuantStudio 7 Pro PCR System (Thermo Fisher Scientific).

#### 6. Additional Analyses

Initially, DDE associations with cognitive performance and biomarker levels were tested in models adjusted only for offspring sex, race, and DDT (continuous ng/mL); then completed education was subsequently added to allow independent examination of its impact on observed associations. To determine whether the adjustment for correlation between siblings was incomplete, we ran sensitivity analyses excluding siblings, of which there were only 8. To determine whether the timing of maternal blood draw during pregnancy affected DDE associations with Aβ 42/40, we adjusted for timing of blood draw (draw within 1st-3rd trimesters vs. post-partum draw) and also ran models including only post-partum blood draws.

1.	Krieger N. Overcoming the absence of socioeconomic data in medical records: validation and application of a census-based methodology. *Am J Public Health*. May 1992;82(5):703-10. doi:10.2105/ajph.82.5.703

2.	van den Berg BJ, Christianson RE, Oechsli FW. The California Child Health and Development Studies of the School of Public Health, University of California at Berkeley. *Paediatr Perinat Epidemiol*. Jul 1988;2(3):265-82. doi:10.1111/j.1365-3016.1988.tb00218.x

3.	Link BG, Susser ES, Factor-Litvak P, et al. Disparities in self-rated health across generations and through the life course. *Soc Sci Med*. Feb 2017;174:17-25. doi:10.1016/j.socscimed.2016.11.035

4.	Sholtz RI, McLaughlin KR, Cirillo PM, et al. Assaying organochlorines in archived serum for a large, long-term cohort: implications of combining assay results from multiple laboratories over time. *Environ Int*. May 2011;37(4):709-14. doi:10.1016/j.envint.2011.01.013

5.	Wechsler D. *WAIS-R Manual: Wechsler Adult Intelligence Scale-revised*. Psychological Corporation; 1981.

6.	Brickman AM, Paul RH, Cohen RA, et al. Category and letter verbal fluency across the adult lifespan: relationship to EEG theta power. *Arch Clin Neuropsychol*. Jul 2005;20(5):561-73. doi:10.1016/j.acn.2004.12.006

7.	Wechsler D. *Wechsler Test of Adult Reading: WTAR*. Psychological Corporation; 2001.
